# A COMPARISON OF TRENDS IN ADIPOSITY AND BLOOD PRESSURE AMONG PEOPLE LIVING WITH HIV USING DOLUTEGRAVIR WITH OTHER ANTIRETROVIRAL THERAPIES IN MWANZA TANZANIA

**DOI:** 10.64898/2026.11.24.26368816

**Authors:** Mayebe Tubeti, Denna Michael, Eveline Konje, Jim Todd, Evangelista Malindisa

## Abstract

**Background:** High blood pressure (BP) is a serious public health concern worldwide, including Tanzania, due to its impact in cardiovascular diseases. Among people living with HIV (PLHIV) high blood pressure has been linked with some anti-retroviral therapy (ART) regimens. Since its introduction in 2018, dolutegravir (DTG) has achieved a high rate of viral suppression compared to previous ART regimens. However, clinical and epidemiological studies have associated DTG with weight gain and other adiposity measures, which may increase the risk of high blood pressure and cardiovascular diseases.

**Method:** Between 1 June 2016 to 31 May 2021 500 adults PLHIV were started on ART, with BP, body mass index (BMI) and waist-to-height ratio (WtHR) measured at initiation and at regular follow-ups. Trends in BP, BMI, and WtHR were described using line graphs. Changes in BP, BMI and WtHR, and 95% confidence intervals (95%CI), associated with DTG or other ART regimes were assessed using mixed-effects linear regression models, with patients as clusters.

**Results:** DTG was significantly associated with increase in BMI (coefficient=0.03, p=0.001, 95%CI=0.02-0.04) after adjusted for age and sex compared to other ART. DTG was not significantly associated with change in systolic BP compared with other ART regimens (coefficient=0.003, p=0.886, 95% CI: −0.04 to 0.05) after adjustment for BMI, age and sex.

**Conclusion:** DTG was associated with greater increases in BMI compared to other ART regimens but after adjusting for BMI was not independently linked to higher systolic blood pressure. The absence of an association between DTG and SBP after adjustment for BMI suggests that differences in adiposity may partly account for the observed crude association.

## Background

High blood pressure is a widespread and significant health concern among adults globally, affecting approximately 1.28 billion people aged 30-79 years (1–3). Many individuals with high blood pressure are unaware of their condition, hindering efforts toward treatment and control (3). High blood pressure is a major risk factor for cardiovascular and renal diseases, including coronary heart disease, heart failure, stroke, peripheral artery disease, and end-stage renal disease, and contributes substantially to premature mortality worldwide (4)(5). High blood pressure is defined as a sustained elevation in blood pressure to ≥140 mmHg systolic or ≥90 mmHg diastolic (4).

In Africa, more than 30% of all adults aged 25 and older have hypertension, with contributing factors including malnutrition, rapid urbanization, and physical inactivity(6–7). A study in Tanzania in 2016, reported 8.0% of adults aged 15 to 55 years had high blood pressure, with higher rates among older adults (30.8%), and females (8.2%) than in males (7.7%). The majority of high blood pressure patients were not aware of their status (8). A study in Morogoro in 2020 showed that 29.3% of adults aged 25 to 64 had hypertension, with 34.3% of them being aware of their condition and 29.9% receiving antihypertensive medications(9) (9–10). In Kilimanjaro region in 2023 hypertension was found in 55.2% of hospital based patients aged 18 to 70+ years (11).

Among people living with HIV (PLHIV), hypertension is common and may be influenced by both traditional cardiovascular risk factors and exposure to antiretroviral therapy (ART) (12)(13). Dolutegravir (DTG), an integrase strand transfer inhibitor (INSTI), provides rapid viral suppression and is recommended by the World Health Organization (WHO) as a first-line treatment for adults with HIV; Tanzania adopted DTG-based regimens as part of first-line HIV treatment in 2019 (14)(15). However, evidence indicates that DTG-based regimens are associated with greater weight gain than some other ART regimens, with greater weight gain reported among women than men (15). Weight gain is associated with an increased risk of hypertension (16). Adiposity can be assessed using measures such as body mass index (BMI), waist circumference, waist-to-height ratio (WtHR), and waist-to-hip ratio (16)(17), In particular, abdominal adiposity has been associated with an increased risk of hypertension (17). Substantial weight gain may increase the risk of hypertension, cardiovascular disease, and other serious non-HIV-related conditions (15)(18)(19).

Moreover, studies among PLHIV receiving DTG-based ART have reported associations between weight gain and increases in blood pressure (18)(20). A recent target-trial emulation using data from the REPRIEVE cohort found that switching to an integrase inhibitor was associated with increased risks of obesity, diabetes, and hypertension, but not major adverse cardiovascular events (21). However, it remains unclear whether DTG is independently associated with increases in blood pressure or whether the observed relationship is primarily related to DTG-associated weight gain.

This study builds on previous research in Mwanza, Tanzania, which reported associations between DTG use and diabetes, insulin resistance, and weight gain (14). Given the potential cardiometabolic consequences of DTG-associated weight gain, there is a need to examine longitudinal trends in adiposity and blood pressure and their associations among PLHIV receiving DTG-based and other ART regimens (22).

## Materials and Methods

This analysis used data from a cohort of adults living with HIV who initiated ART at three public HIV clinics in Mwanza, Tanzania, and were followed for four years from 1 June 2016 to 31 may 2021. Written informed consent was obtained from all participants(23). Participants were excluded if they lived outside Mwanza, were unable or unwilling to provide informed consent, had previously received ART, did not have access to a phone, were younger than 18 years, or had an estimated survival prognosis of less than one year.

At the first visit, all participants were screened for cardiovascular conditions. Data on sociodemographic and lifestyle characteristics, including age, sex, education level, tobacco use, alcohol use, physical activity, dietary patterns, stress, and sleep, were collected using a structured questionnaire. Anthropometric measurements and blood pressure were assessed at baseline and at subsequent clinic visits. Data were double-entered into Open Clinica software Two measures of adiposity were included in this analysis: BMI and WtHR. BMI was calculated as weight in kilograms divided by height in metres squared (kg/m²), while WtHR was calculated as waist circumference divided by height. These measures were derived during analysis from the anthropometric measurements collected during the study.

Blood pressure was measured at each study visit using an OMRON HBP-1300 professional blood pressure monitor, following the standardized blood pressure measurement procedure of the National Health and Nutrition Examination Survey (NHANES). Participants rested for 5 minutes in a seated position with their back supported, legs uncrossed, and feet flat on the floor. Three successive blood pressure measurements were taken on the right arm at 60-second intervals.

The adiposity variables (BMI, WtHR) and both systolic blood pressure (SBP) and diastolic blood pressure (DBP) were analysed as continuous variables and summarized using means and standard deviations across sociodemographic characteristics. Box plots were used to compare adiposity and blood pressure across ART regimens. Histogram were used to assess the distribution of the data, and line graphs were used to show trends in mean values over time.

Mixed-effects models, with individuals as clusters, were used to analyse the four outcomes over time, with the null hypothesis that there was no difference between DTG and other ART regimens. The adiposity outcomes were adjusted for age and sex, while SBP and DBP were further adjusted separately for BMI and WtHR. STATA version 15 was used for all analyses.

This secondary analysis included 494 people living with HIV who contributed 3,155 SBP observations over a maximum follow-up period of 48 months. Power was evaluated in relation to the longitudinal mixed-effects design, accounting for repeated measurements within participants rather than treating observations as independent. Participants had up to 10 scheduled measurements at 0, 3, 6, 12, 18, 24, 30, 36, 42, and 48 months, with an average of 6.2 observed SBP measurements per participant in the mixed-effects analysis. The model included time, DTG status, their interaction, and participant-specific random intercepts and slopes. The estimated residual variance was 64.87 (SD 8.05 mmHg), with a participant-level random-intercept variance of 167.65 and random-slope variance of 0.0234. The approximate within-participant correlation was 0.73. At a two-sided significance level of 5%, the estimated DTG-by-time effect was −0.120 mmHg per month, equivalent to −1.44 mmHg per year. Based on the observed standard error of the longitudinal interaction effect, the study could detect an annual difference of approximately 0.93 mmHg with 80% power and 1.07 mmHg with 90% power. The observed annual difference of 1.44 mmHg corresponds to an approximate two-sided power of 99.2%

## Results

### Descriptive characteristics of subjects in the cohort

Six participants who never initiated ART were excluded, leaving 494 participants in the analysis, contributing a total of 3,155 visits (Table 1). Of the 3,155 visits, 1,108 occurred while participants were receiving DTG-based ART and 1,947 while receiving other ART regimens. Among the 494 participants, 68 received only other ART regimens, 57 received only DTG-based ART, and 369 initially received other ART regimens before switching to DTG-based ART. DTG exposure was treated as a time-varying exposure. At each follow-up visit, participants were classified according to the ART regimen they were receiving at that time. For participants who switched from a non-DTG ART regimen to a DTG-based regimen during follow-up, observations before the switch were classified as pre-DTG exposure, whereas observations after the switch were classified as post-DTG exposure. There were 151 (30.57%) male and 343 (69.43%) female participants, with similar proportions of visits contributed by males and females while receiving DTG-based and other ART regimens (Table 1).

**Table 1:** Description of 494 HIV patient’s cohort on ART by DTG status and number of visits.

| ART Status | Total | Number of patients |  | Number of visits |  |
|---|---|---|---|---|---|
|  |  | Other ART | On DTG | Other ART | On DTG |
| Both other ART and DTG | 369 | 369 | 369 | 1,732 | 927 |
| Started DTG, no other ART | 57 | - | 57 | 0 | 181 |
| On other ART, no DTG | 68 | 68 | - | 215 | 0 |
| Total | 494 | 431 | 426 | 1,947 | 1,108 |

### Description of the trends in adiposity and blood pressure

The follow-up visits, with mean values for adiposity and blood pressure measures among participants receiving other ART before switching to DTG and those receiving DTG, are shown in Table 2. A total of 233 (47%) and 162 (33%) participants were followed for 24 months for other Art and DTG respectively. Table 2 shows the trends in the four outcome variables over time. This trend has presented in by group separately in supplementary table 2, and 3.

**Table 2.** Follow-up and mean adiposity and blood pressure measures among 494 participants receiving DTG-based and other ART regime.

| **Time (months)** | **ART regimen** | **n** | **SBP, mean (SD) (mmHg)** | **DBP, mean (SD) (mmHg)** | **BMI, mean (SD) (kg/m<sup>2</sup>)** | **WtHR, mean (SD)** |
|---|---|---|---|---|---|---|
| 0 | DTG | 53 | 119 (17.37) | 74 (11.95) | 22.17 (4.56) | 0.48 (0.06) |
| 0 | Other ART | 386 | 113 (16.46) | 75 (10.91) | 22.37 (4.72) | 0.48 (0.06) |
| 3 | DTG | 72 | 120 (17.65) | 74 (12.23) | 22.68 (4.43) | 0.48 (0.07) |
| 3 | Other ART | 338 | 112 (14.56) | 74 (9.76) | 23.22 (4.94) | 0.50 (0.07) |
| 6 | DTG | 98 | 119 (16.54) | 73 (11.63) | 22.99 (4.13) | 0.49 (0.06) |
| 6 | Other ART | 326 | 113 (15.17) | 74 (10.03) | 23.32 (4.98) | 0.51 (0.06) |
| 12 | DTG | 101 | 118 (16.08) | 73 (11.87) | 23.18 (4.25) | 0.49 (0.07) |
| 12 | Other ART | 257 | 116 (14.07) | 74 (10.13) | 23.65 (5.04) | 0.51 (0.06) |
| 18 | DTG | 41 | 116 (14.51) | 73 (9.16) | 23.86 (4.59) | 0.51 (0.07) |
| 18 | Other ART | 251 | 117 (14.37) | 74 (9.44) | 23.72 (5.31) | 0.52 (0.07) |
| 24 | DTG | 162 | 116 (14.70) | 70 (10.67) | 23.86 (4.33) | 0.49 (0.06) |
| 24 | Other ART | 233 | 117 (14.15) | 73 (9.87) | 23.61 (5.42) | 0.51 (0.07) |
| 30 | DTG | 122 | 114 (13.52) | 71 (10.48) | 23.03 (5.37) | 0.49 (0.07) |
| 30 | Other ART | 114 | 115 (15.53) | 72 (11.83) | 24.30 (5.47) | 0.51 (0.07) |
| 36 | DTG | 172 | 114 (14.42) | 72 (10.95) | 23.98 (5.68) | 0.51 (0.07) |
| 36 | Other ART | 26 | 115 (12.81) | 71 (8.71) | 23.51 (4.25) | 0.49 (0.05) |
| 42 | DTG | 61 | 114 (17.01) | 72 (12.10) | 24.13 (4.52) | 0.52 (0.06) |
| 42 | Other ART | 9 | 115 (8.57) | 70 (7.22) | 22.66 (3.91) | 0.48 (0.05) |
| 48 | DTG | 226 | 117 (15.07) | 72 (11.59) | 24.92 (5.82) | 0.51 (0.07) |
| 48 | Other ART | 7 | 110 (10.65) | 68 (9.56) | 21.42 (3.99) | 0.47 (0.07) |

### Analysis of the effect of dolutegravir on adiposity over time

In the mixed-effects model accounting for clustering by participant, there was no significant difference in BMI between DTG and other ART regimens at baseline. BMI increased over time among participants receiving both DTG and other ART regimens (Table 3). The estimated monthly increase was 0.03 kg/m² (95% CI: 0.03–0.04) among participants receiving DTG and 0.02 kg/m² (95% CI: 0.02–0.03) among those receiving other ART regimens. The difference in the rates of BMI increase between the two groups was statistically significant (p<0.009). After adjustment for age and sex, the monthly increase in BMI remained similar to the crude estimates in both the DTG and other ART groups (p<0.001 for both groups) (Table 3). The mixed-effects model accounted for substantial between-participant variation in BMI (variance = 20.22) and variation in monthly BMI change within participants over time (variance = 1.25)

**Table 3:**
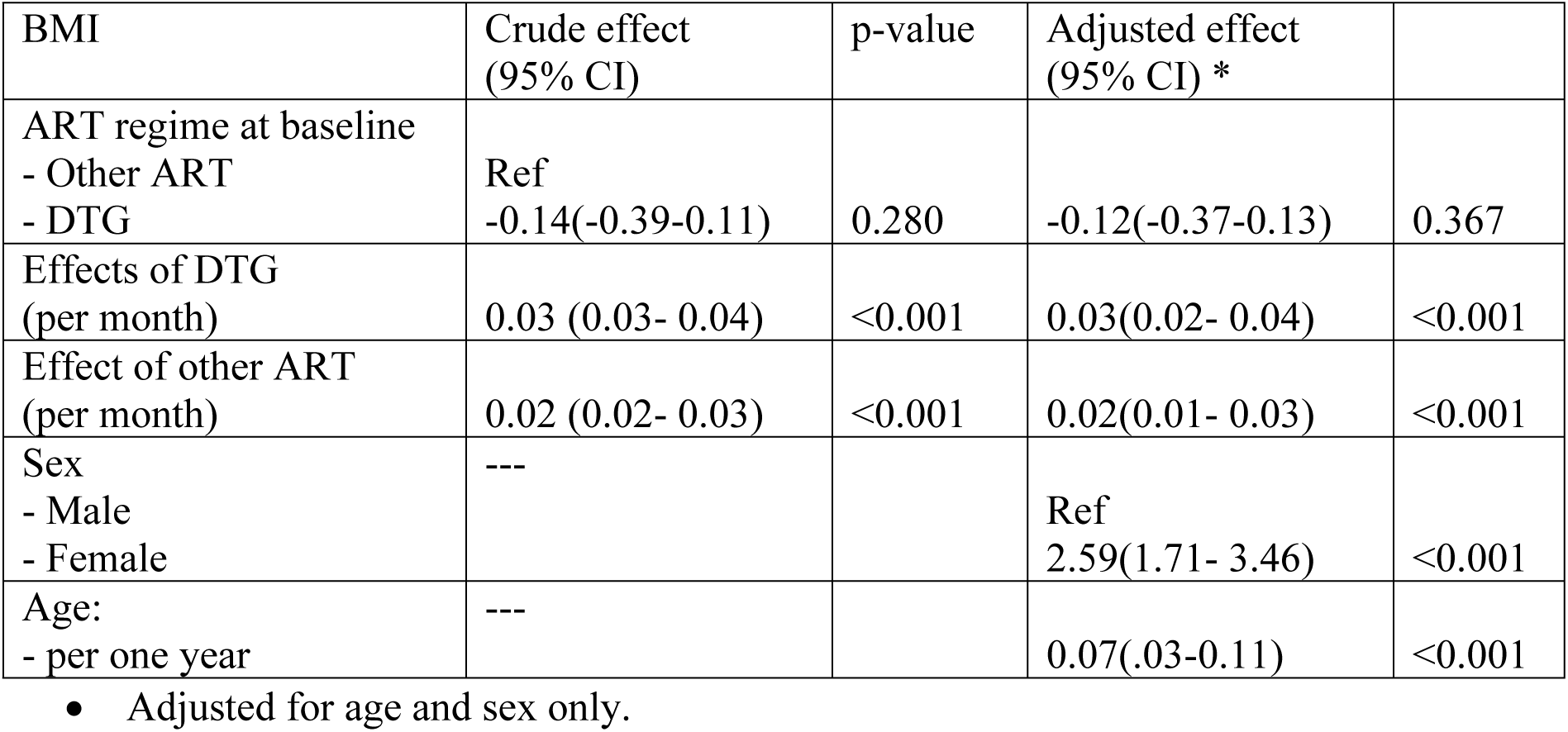
Effect of DTG on Body Mass Index using mixed effect model accounting for differences between individual patients.

| BMI | Crude effect (95% CI) | p-value | Adjusted effect (95% CI) * |  |
|---|---|---|---|---|
| ART regime at baseline |  |  |  |  |
| - Other ART | Ref |  |  |  |
| - DTG | -0.14(-0.39-0.11) | 0.280 | -0.12(-0.37-0.13) | 0.367 |
| Effects of DTG (per month) | 0.03 (0.03- 0.04) | <0.001 | 0.03(0.02- 0.04) | <0.001 |
| Effect of other ART (per month) | 0.02 (0.02- 0.03) | <0.001 | 0.02(0.01- 0.03) | <0.001 |
| Sex | --- |  |  |  |
| - Male |  |  | Ref |  |
| - Female |  |  | 2.59(1.71- 3.46) | <0.001 |
| Age: | --- |  |  |  |
| - per one year |  |  | 0.07(.03-0.11) | <0.001 |

• Adjusted for age and sex only.

For waist-to-height ratio (WtHR), there was no significant increase over time among participants receiving DTG (0.008 per month, p=0.380), whereas WtHR increased significantly among those receiving other ART regimens (0.02 per month, p<0.001) in the crude model (Index Table 4). After adjustment for age and sex, there was no significant change in WtHR over time in either the DTG group (−0.008 per month, p=0.359) or the other ART group (0.01 per month, p=0.085) (supplementary Table 4). The mixed-effects model accounted for substantial between-participant variation in WtHR (variance = 32.13) and within-participant variation over time (residual variance = 6.65) (supplementary Table 4).

**Table 4.** Effect of DTG on SBP adjusted for adiposity (BMI and WtHR) over time.

| **SBP** | **Crude effect for SBP (95% CI)** | **P** | **Effect adjusted for BMI (95%CI)** | **P** | **Adjusted effect for WtHR** | **P** |
|---|---|---|---|---|---|---|
| ART regime<br>-Other ART<br>-DTG | Ref<br>0.30(-1.28-1.89) | 0.708 | Ref<br>0.21(-1.34- 1.75) | 0.790 | 0.19(-1.36- 1.74) | 0.809 |
| Effects of DTG (over time) | 0.05(0.001-0.10) | 0.046 | 0.003(-0.04- 0.05) | 0.886 | 0.02(-0.02- 0.07) | 0.320 |
| Effect of other ART (over time) | 0.17(0.13-0.21) | < 0.001 | 0.13(0.09- 0.17) | <0.001 | 0.13(0.09- 0.17) | <0.001 |
| BMI (per unit increase) | --- |  | 0.89(0.73- 1.06) | <0.001 | --- |  |
| WtHR | --- |  | --- |  | 0.43(0.33- 0.53) | <0.001 |
| Sex<br>-Male<br>-Female | --- |  | Ref<br>-9.01(-11.39- -6.62) | <0.001 | -8.88(-11.32- -6.44) | <0.001 |
| Age (per year increase) | --- |  | 0.30(0.19- 0.41) | <0.001 | 0.31(0.19-0.42) | <0.001 |

• Adjusted for age, sex and adiposity (BMI or WtHR)

### Effect of dolutegravir on blood pressure over time

After adjustment for age, sex and BMI, dolutegravir (DTG) was not significantly associated with baseline systolic blood pressure (SBP) compared with other ART regimens (coefficient = 0.21 mmHg, p=0.790, 95% CI: −1.34 to 1.75). Over time, SBP increased slightly among participants receiving DTG-based ART, but the increase was not statistically significant (coefficient = 0.003 mmHg/month, p=0.886, 95% CI: −0.04 to 0.05). In contrast, SBP increased significantly over time among participants receiving other ART regimens (coefficient = 0.13 mmHg/month, p<0.001, 95% CI: 0.09 to 0.17). The rate decrease change in SBP differed significantly between the DTG and other ART groups (p < 0.001) (Table 4).

After adjustment for age, sex and WtHR, DTG was also not significantly associated with baseline SBP compared with other ART regimens (coefficient = 0.19 mmHg, p=0.809, 95% CI: −1.36 to 1.74). Over time, the increase in SBP among participants receiving DTG-based ART remained statistically non-significant (coefficient = 0.02 mmHg/month, p=0.320, 95% CI: −0.02 to 0.07), whereas SBP increased significantly among those receiving other ART regimens (coefficient = 0.13 mmHg/month, p<0.001, 95% CI: 0.09 to 0.17) (Table 4). Importantly, the rate of change in SBP differed significantly between the two ART groups (difference in slopes = −0.11 mmHg/month, p < 0.001, 95% CI: −0.16 to −0.06), indicating that SBP increased at a significantly slower rate among participants receiving DTG-based ART than among those receiving other ART regimens. The random-effects model showed greater between-participant variation in SBP (variance = 133) than within-participant variation over time (residual variance = 64.34).

After adjustment for age, sex, and BMI, dolutegravir-based ART was significantly associated with lower diastolic blood pressure (DBP) at baseline compared with other ART regimens (coefficient = −1.32 mmHg, 95% CI: −2.49 to −0.15; p=0.038). Over time, DBP decreased significantly among participants receiving DTG-based ART (coefficient = −0.09 mmHg/month, 95% CI: −0.12 to −0.05), while DBP also decreased among participants receiving other ART regimens (coefficient = −0.12 mmHg/month, 95% CI: −0.15 to −0.08; p<0.001) after adjustment for age, sex, and BMI andthe difference in the rate of change in DBP between the DTG and other ART groups was not statistically significant (p= 0.167).

When WtHR was included instead of BMI, DTG remained significantly associated with lower DBP at baseline compared with other ART regimens (coefficient = −1.30 mmHg, 95% CI: −2.48 to −0.13; p=0.030). Over time, DBP decreased among participants receiving DTG-based ART (coefficient = −0.08 mmHg/month, 95% CI: −0.11 to −0.03) and among those receiving other ART regimens (coefficient = −0.11 mmHg/month, 95% CI: −0.15 to −0.08) after adjustment for age, sex, and WtHR. However, the rate of change in DBP did not differ significantly between the DTG and other ART groups (p = 0.059). The random-effects model showed greater between-participant variation in DBP (variance = 58.06) than within-participant variation over time (residual variance = 38.02) (Table 5)

**Table 5.**
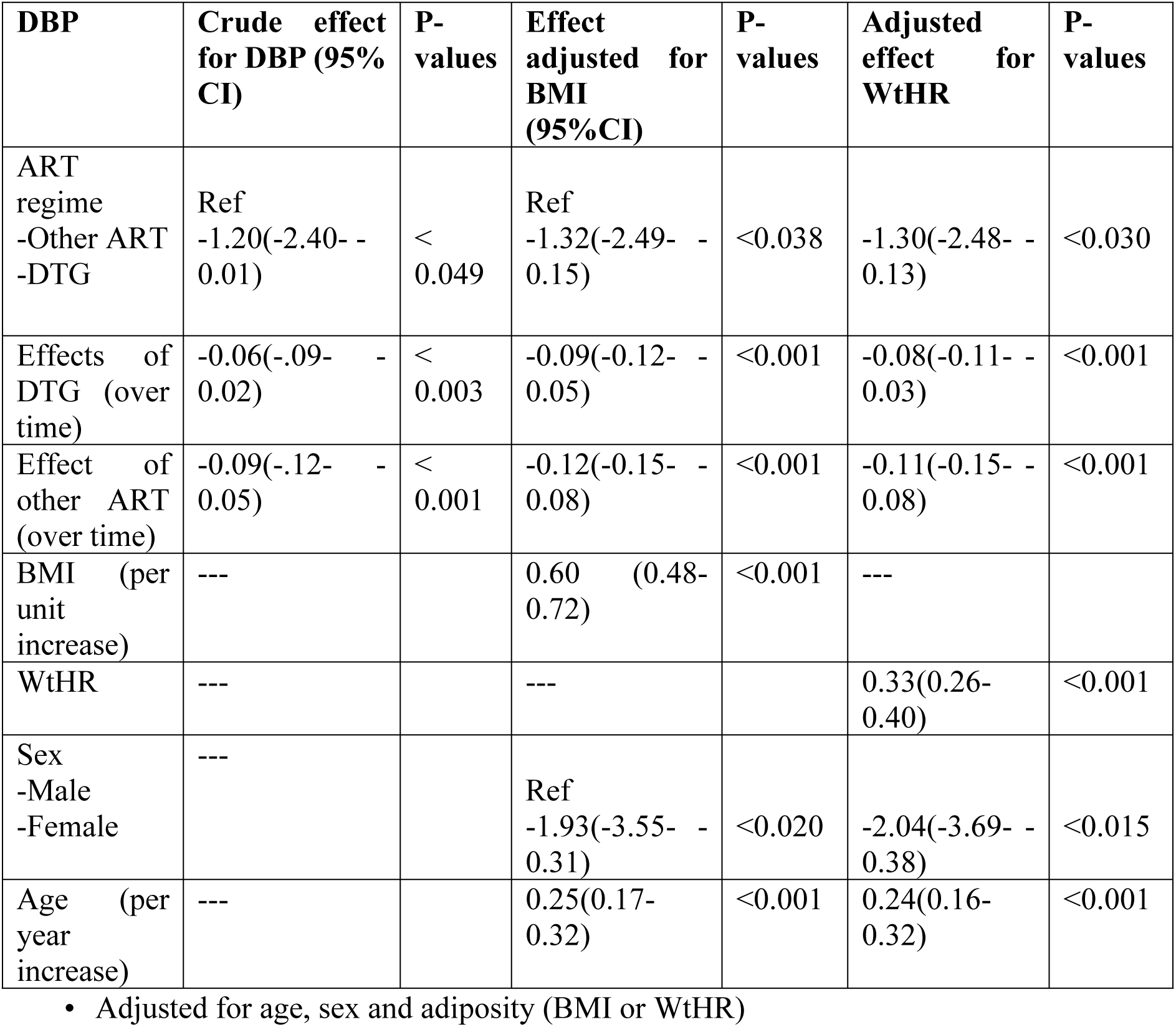
Effect of DTG on DBP adjusted for adiposity (BMI and WtHR) over time.

| **DBP** | **Crude effect for DBP (95% CI)** | **P-values** | **Effect adjusted for BMI (95%CI)** | **P-values** | **Adjusted effect for WtHR** | **P-values** |
|---|---|---|---|---|---|---|
| ART regime<br>-Other ART<br>-DTG | Ref<br>-1.20(-2.40- -0.01) | < 0.049 | Ref<br>-1.32(-2.49- -0.15) | <0.038 | -1.30(-2.48- -0.13) | <0.030 |
| Effects of DTG (over time) | -0.06(-.09- -0.02) | < 0.003 | -0.09(-0.12- -0.05) | <0.001 | -0.08(-0.11- -0.03) | <0.001 |
| Effect of other ART (over time) | -0.09(-.12- -0.05) | < 0.001 | -0.12(-0.15- -0.08) | <0.001 | -0.11(-0.15- -0.08) | <0.001 |
| BMI (per unit increase) | --- |  | 0.60 (0.48-0.72) | <0.001 | --- |  |
| WtHR | --- |  | --- |  | 0.33(0.26-0.40) | <0.001 |
| Sex<br>-Male<br>-Female | --- |  | Ref<br>-1.93(-3.55- -0.31) | <0.020 | -2.04(-3.69- -0.38) | <0.015 |
| Age (per year increase) | --- |  | 0.25(0.17-0.32) | <0.001 | 0.24(0.16-0.32) | <0.001 |

• Adjusted for age, sex and adiposity (BMI or WtHR)

## Discussion

This longitudinal study found that participants receiving dolutegravir (DTG)-based antiretroviral therapy (ART) had a greater increase in body mass index (BMI) over time than those receiving other ART regimens. BMI increased in both groups, but the rate of increase was higher among participants receiving DTG. In contrast, there was no clear evidence of an increase in waist-to-height ratio (WtHR) associated with DTG after adjustment for age and sex. Systolic blood pressure (SBP) increased over time among participants receiving DTG in the crude analysis, but this association was substantially attenuated after adjustment for BMI or WtHR. These findings suggest that adiposity may be an important factor in the relationship between DTG-based ART and changes in SBP, rather than providing evidence of an independent association between DTG and SBP.

The greater increase in BMI among participants receiving DTG is consistent with previous evidence linking DTG and other integrase inhibitor-based regimens with greater weight gain than some older ART regimens. Studies from sub-Saharan Africa have reported weight and BMI increases following DTG initiation or switching, with greater increases observed in some groups, particularly women (15) (24). The present findings therefore add longitudinal evidence from Tanzania to the growing literature on DTG-associated weight gain. However, weight gain following ART initiation is likely to be multifactorial and may reflect recovery from HIV-related illness, changes in appetite and energy balance, baseline nutritional status, sex, age and other characteristics in addition to the specific ART regimen(25). Thus, the greater BMI increase observed with DTG should be interpreted as an association rather than evidence of a direct pharmacological effect.

The relationship between adiposity and SBP is particularly relevant to interpreting our findings. BMI was positively associated with SBP, while the crude increase in SBP among participants receiving DTG was attenuated to essentially no association after adjustment for BMI. A similar pattern was reported in the ADVANCE trial, where changes in blood pressure were associated with weight gain rather than ART regimen after adjustment for BMI (18). This consistency suggests that changes in adiposity may contribute importantly to changes in blood pressure observed during ART. However, BMI was included as an adjustment variable in the present analysis and no formal mediation analysis was performed. Therefore, our findings do not establish that BMI mediates an effect of DTG on SBP; rather, they indicate that the observed DTG–SBP association was not independent of adiposity.

Evidence on the relationship between DTG and blood pressure remains mixed. In the present study, the crude increase in SBP among DTG users was small and was no longer statistically significant after adjustment for adiposity. This is consistent with the ADVANCE findings (18), but differs from the original D2EFT analysis, which reported greater increases in SBP and DBP in a DTG-containing treatment arm compared with the comparator (26). In contrast, the NEAT-022 study found no significant difference in blood pressure changes or incident hypertension following switching to DTG compared with continued protease inhibitor-based therapy (27). More recently, the REPRIEVE target-trial emulation found that switching to an integrase inhibitor was associated with increased risks of obesity, diabetes and hypertension, although not major adverse cardiovascular events (21). Most participants switching to an integrase inhibitor initiated DTG-based therapy. Together, these findings suggest that the relationship between DTG and cardiometabolic outcomes is complex and may be influenced by weight gain, baseline cardiovascular risk, comparator regimen and duration of follow-up. Importantly, our study assessed longitudinal SBP and DBP rather than incident hypertension; therefore, the absence of an independent association with SBP should not be interpreted as evidence that DTG has no effect on hypertension risk.

The decrease in DBP observed over time among participants receiving both DTG and other ART regimens should also be interpreted cautiously. Although DBP declined significantly within both groups, this does not by itself demonstrate that DTG was associated with a greater reduction than other ART. The direction of these findings also differs from some previous studies reporting increases in DBP following DTG exposure (26) (27). Factors common to longitudinal HIV care, changes following ART initiation, regression toward the mean and other time-varying characteristics may contribute to the observed pattern. Similarly, the absence of a significant longitudinal increase in WtHR after adjustment suggests that the greater increase in BMI was not accompanied by a clearly detectable increase in this measure of central adiposity. Because BMI and WtHR capture different aspects of adiposity, future studies incorporating direct measures of body composition and fat distribution may help clarify the nature of DTG-associated weight gain.

This study has several strengths, including the use of longitudinal data with repeated measurements of adiposity and blood pressure and mixed-effects models that accounted for repeated observations within participants. The inclusion of both BMI and WtHR also allowed overall and central adiposity to be considered separately. However, several limitations warrant consideration. First, this was a secondary observational analysis and therefore cannot establish causality or exclude residual confounding. Second, most participants who received DTG had initially received another ART regimen and subsequently switched to DTG. This treatment switching creates a time-varying exposure structure and may introduce confounding related to treatment selection and calendar time. Third, BMI and WtHR are indirect measures of body composition and cannot distinguish changes in fat mass from lean mass. Fourth, fewer participants contributed observations at later follow-up visits, which may reduce precision in estimating longer-term trajectories. Finally, adjustment for BMI or WtHR changes the estimand and may account for part of a potential pathway between ART exposure and blood pressure.

From a clinical perspective, the findings support continued monitoring of weight and cardiometabolic risk among people living with HIV receiving DTG-based ART. The absence of an independent association between DTG and increasing SBP after adjustment for adiposity is reassuring, while the greater increase in BMI highlights the importance of incorporating simple measures of adiposity and blood pressure into long-term HIV care. These findings do not suggest that DTG-based ART should be avoided, but rather that the potential cardiometabolic consequences of weight gain should be recognized and monitored. Prospective studies with longer follow-up, more detailed measures of body composition and metabolic risk, and appropriate methods for handling treatment switching are needed to determine whether DTG-associated weight gain translates into clinically meaningful increases in hypertension and cardiovascular risk.

Overall, DTG-based ART was associated with a greater increase in BMI over time but was not independently associated with increasing SBP after accounting for adiposity. The findings therefore support a potential role of adiposity in the relationship between DTG-based ART and blood pressure, while not demonstrating a direct or causal effect of DTG on SBP. The contrasting DBP pattern and the absence of a clear increase in WtHR further emphasize the need for cautious interpretation and continued investigation of the long-term cardiometabolic effects of DTG-based ART.

### Conclusion and Recommendations

DTG-based ART was associated with a greater increase in adiposity over time than other ART regimens, which was measured by body mass index and waist to height ratio in this study. However, DTG was not independently associated with increase systolic blood pressure after adjustment for adiposity. These findings suggest that adiposity may be an important contributor to the relationship between DTG use and blood pressure. Routine monitoring of weight, adiposity and blood pressure should therefore be integrated into long-term HIV care for people receiving DTG-based ART. Further longitudinal studies with longer follow-up and more direct measures of body composition are recommended to clarify whether DTG-associated weight gain contributes to subsequent hypertension and cardiovascular risk.

## Declarations Ethics approval

In carrying the study, the ethical clearance was sought to CUHAS/BMC joint ethics and review committee for increasing research credibility and integrity. The ethical clearance number CREC/933/2025 was given from the committee.

## Consent for publication

Not applicable

## Availability of data and materials

The data used and/or analysed during this study are not publicly available due to ethical and confidentiality considerations involving human subjects. However, the datasets may be made available from the corresponding author upon reasonable request and with permission from the relevant ethics committee.

## Authors’ contributions

Mayebe Tubeti conducted the data analysis and manuscript preparation. Dr. Evangelista Malindisa provided guidance, editing and revised manuscript. Prof. Jim Todd offered guidance on data analysis, editing and revised the manuscript, Dr. Konje provided insightful suggestions to improve the quality and revised manuscript and Dr. Denna Michael involved in editing and revised the manuscript.

## Data Availability

Yes, All relevant data from this study will be made available upon study completion.

## Acknowledgements

We would like to extend our heartfelt thanks to CUHAS and the Department of Epidemiology and Biostatistics for providing the necessary academic and material support that sustained this research from its inception to completion. We are also deeply grateful to Professor Rob Peck for his valuable guidance, support, and contributions throughout this research

## Supplementary tables

**Supplementary Table 2:** Follow up of 494 subjects and mean adiposity and blood pressure for patients on other ART at each follow up visit.

| Time points in months | Outcomes |  | Adiposity |  | Total patients |
|---|---|---|---|---|---|
|  | BP mean (sd) |  | BMI<br>Mean(sd)<br>(kg/m<sup>2</sup>) | WtHR<br>Mean(sd) |  |
|  | SBP (mmHg) | DBP (mmHg) |  |  |  |
| 0 | 113 (16.46) | 75(10.91) | 22.37 (4.72) | 0.48(0.06) | 386 |
| 3 | 112(14.56) | 74(9.76) | 23.22(4.94) | 0.50(0.07) | 338 |
| 6 | 113(15.17) | 74(10.03) | 23.32(4.98) | 0.51(0.06) | 326 |
| 12 | 116(14.07) | 74(10.13) | 23.65(5.04) | 0.51(0.06) | 257 |
| 18 | 117(14.37) | 74(9.44) | 23.72(5.31) | 0.52(0.07) | 251 |
| 24 | 117(14.15) | 73(9.87) | 23.61(5.42) | 0.51(0.07) | 233 |
| 30 | 115(15.53) | 72(11.83) | 24.30(5.47) | 0.51(0.07) | 114 |
| 36 | 115(12.81) | 71(8.71) | 23.51(4.25) | 0.49(0.05) | 26 |
| 42 | 115(8.57) | 70(7.22) | 22.66(3.91) | 0.48(0.05) | 9 |
| 48 | 110(10.65) | 68(9.56) | 21.42(3.99) | 0.47(0.07) | 7 |

**Supplementary Table 3:**
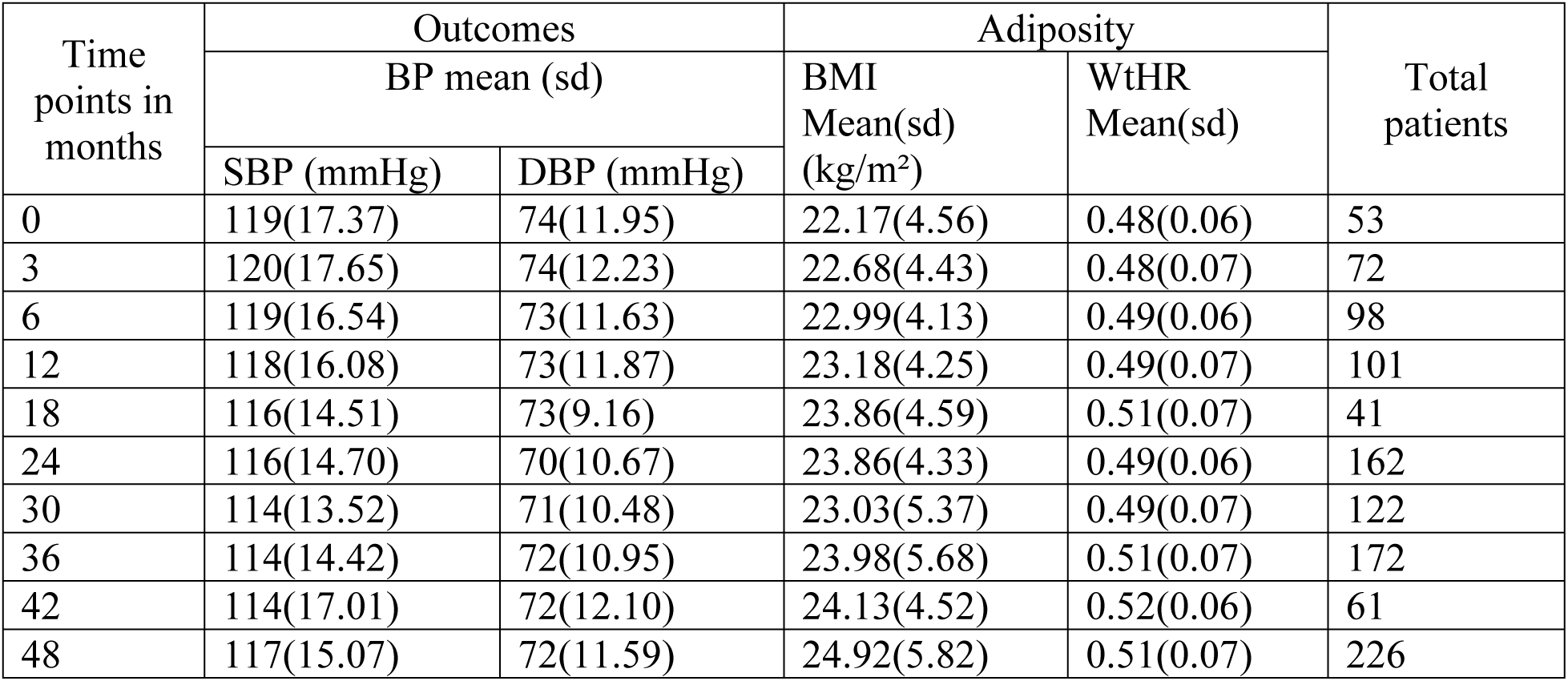
Follow up of 494 subjects and mean adiposity and blood pressure for patients on DTG at each follow up visit.

| Time points in months | Outcomes |  | Adiposity |  | Total patients |
|---|---|---|---|---|---|
|  | BP mean (sd) |  | BMI<br>Mean(sd)<br>(kg/m²) | WtHR<br>Mean(sd) |  |
|  | SBP (mmHg) | DBP (mmHg) |  |  |  |
| 0 | 119(17.37) | 74(11.95) | 22.17(4.56) | 0.48(0.06) | 53 |
| 3 | 120(17.65) | 74(12.23) | 22.68(4.43) | 0.48(0.07) | 72 |
| 6 | 119(16.54) | 73(11.63) | 22.99(4.13) | 0.49(0.06) | 98 |
| 12 | 118(16.08) | 73(11.87) | 23.18(4.25) | 0.49(0.07) | 101 |
| 18 | 116(14.51) | 73(9.16) | 23.86(4.59) | 0.51(0.07) | 41 |
| 24 | 116(14.70) | 70(10.67) | 23.86(4.33) | 0.49(0.06) | 162 |
| 30 | 114(13.52) | 71(10.48) | 23.03(5.37) | 0.49(0.07) | 122 |
| 36 | 114(14.42) | 72(10.95) | 23.98(5.68) | 0.51(0.07) | 172 |
| 42 | 114(17.01) | 72(12.10) | 24.13(4.52) | 0.52(0.06) | 61 |
| 48 | 117(15.07) | 72(11.59) | 24.92(5.82) | 0.51(0.07) | 226 |

**Supplementary Table 4:** Effect of DTG on Waist to Height Ratio using mixed effect model accounting for differences between individual patients.

| Waist to height ratio | Crude effect (95% CI) | p-value | Adjusted effect (95% CI) * | p-value |
|---|---|---|---|---|
| ART regime at baseline<br>- Other ART<br>- DTG | Ref<br>0.01(-0.52- 0.55) | 0.958 | 0.06(-0.47- 0.59) | 0.823 |
| Effects of DTG (per month) | 0.008(-0.01- 0.02) | 0.380 | -0.008( -0.03-0.009) | 0.359 |
| Effect of other ART (per month) | 0.02 (0.01- 0.04) | <0.001 | 0.01 (-0.002- 0.03) | 0.085 |
| Sex<br>- Male<br>- Female | --- |  | Ref<br>5.09 (3.96- 6.23) | <0.001 |
| Age:<br>- per one year | --- |  | 0.18(0.12- 0.23) | <0.001 |

Adjusted for age and sex only.

## List of abbreviations

ART: Antiretroviral Therapy
BMI: Body mass index
BP: blood pressure
DBP: Diastolic blood pressure
DTG: Dolutegravir
HIV: Human Immunodeficiency Virus
INSTIs: Integrase strand transfer inhibitors
NNRTIs: non-nucleoside reverse transcriptase inhibitors
NRTIs: Nucleotide-transcriptase inhibitors
PIs: Protease inhibitors
PLWH: People living with HIV
SBP: Systolic blood pressure
WHO: World Health Organization
WtHR: Waist-to-Height Ratio

## References

1. WHO. More than 700 million people with untreated hypertension. 2021; Available from: https://www.who.int/news/item/25-08-2021-more-than-700-million-people-with-untreated-hypertension

2. Bin Zhou, Rodrigo M Carrillo et al. Articles Worldwide trends in hypertension prevalence and progress in treatment and control from 1990 to 2019: a pooled analysis of 1201 population-representative studies with 104 million participants. 2021;398.

3. WHO. Hypertension. 2023; Available from: https://www.who.int/news-room/fact-sheets/detail/hypertension

4. Mills KT, Stefanescu A, He J. The global epidemiology of hypertension. 2021;16(4):223–37.

5. Fuchs FD, Whelton PK. High Blood Pressure and Cardiovascular Disease. Hypertension. 2020;75(2):285–92.

6. World Health Organization. Hypertension [Internet]. 2023. Available from: https://www.who.int/news-room/fact-sheets/detail/hypertension

7. Kofi W, Id B, Reilly ST, Moses J, Aheto K, Zucchelli E. Hypertension in older adults in Africa: A systematic review and meta-analysis. 2019;1–25.

8. Mosha NR, Mahande M, Juma A, Mboya I, Peck R, Urassa M, et al. Prevalence, awareness and factors associated with hypertension in North West Tanzania. Glob Health Action [Internet]. 2017;10(1). Available from: 10.1080/16549716.2017.1321279

9. Muhihi AJ, Anaeli A, Mpembeni RNM, Sunguya BF, Leyna G, Kakoko D, et al. Prevalence, Awareness, Treatment, and Control of Hypertension among Young and Middle-Aged Adults: Results from a Community-Based Survey in Rural Tanzania. Int J Hypertens. 2020;2020.

10. Nsanya MK, Ayieko P, Hashim R, Mgema E, Fitzgerald D, Kapiga S, et al. Sustained high blood pressure and 24-h ambulatory blood pressure monitoring in Tanzanian adolescents. Sci Rep [Internet]. 2021;11(1):1–9. Available from: 10.1038/s41598-021-87996-0

11. Mwimo J, Kimondo FC, Mboya IB. Prevalence of hypertension and associated factors among diabetic patients in Kilimanjaro region, northern Tanzania: A hospital-based cross-sectional study. Clin Epidemiol Glob Heal [Internet]. 2023;23(June):101387. Available from: 10.1016/j.cegh.2023.101387

12. Rezende AC, Souza LG, Jardim TV, Perillo NB, Araújo YCL, De Souza SG, et al. Is waist-to-height ratio the best predictive indicator of hypertension incidence? A cohort study. BMC Public Health. 2018;18(1):1–11.

13. Fiseha T, Belete AG, Dereje H, Dires A. Hypertension in HIV-Infected Patients Receiving Antiretroviral Therapy in Northeast Ethiopia. Int J Hypertens. 2019;2019.

14. Bangalee A, Hanley S, Bangalee V. Dolutegravir as first-line antiretroviral therapy in South Africa: Beware the one-size-fits-all approach. South African Med J. 2022;112(10):787–90.

15. Brennan AT, Nattey C, Kileel EM, Rosen S, Maskew M, Stokes AC, et al. Change in body weight and risk of hypertension after switching from efavirenz to dolutegravir in adults living with HIV: evidence from routine care in Johannesburg, South Africa. eClinicalMedicine [Internet]. 2023;57:101836. Available from: 10.1016/j.eclinm.2023.101836

16. Zhou B, Bennett JE, Wickham AP, Singleton RK, Mishra A, Carrillo-Larco RM, et al. General and abdominal adiposity and hypertension in eight world regions: a pooled analysis of 837 population-based studies with 7·5 million participants. Lancet. 2024;404(10455):851–63.

17. Dzudie A, Njedock N, Boombhi J, Ba H, Amougou SN, Kamdem F, et al. Association between measures of adiposity and blood pressure levels in adult Cameroonians. Heal Sci Reports. 2021;4(2).

18. Manne-Goehler J, Fabian J, Sokhela S, Akpomiemie G, Rahim N, Lalla-Edward ST, et al. Blood pressure increases are associated with weight gain and not antiretroviral regimen or kidney function: a secondary analysis from the ADVANCE trial in South Africa. J Int AIDS Soc. 2024;27(7):1–9.

19. Verkouter I, Noordam R, Loh NY, Van Dijk KW, Zock PL, Mook-Kanamori DO, et al. The Relation between Adult Weight Gain, Adipocyte Volume, and the Metabolic Profile at Middle Age. J Clin Endocrinol Metab. 2021;106(11):E4438–47.

20. Shamu T, Egger M, Mudzviti T, Chimbetete C, Manasa J, Anderegg N. Body weight and blood pressure changes on dolutegravir-, efavirenz- or atazanavir-based antiretroviral therapy in Zimbabwe: a longitudinal study. J Int AIDS Soc. 2024;27(2):1–12.

21. Kileel EM, Malvestutto CD, Zanni M V, Davies-smith E, Fichtenbaum CJ, Lake JE, et al. Articles Risk of obesity, diabetes, hypertension, and major adverse cardiovascular events after a switch to an integrase inhibitor: a target trial emulation in REPRIEVE. 2026;3018(25):1–10.

22. Kisigo GA, Barongo E, Issarow B, Cichowitz C, Wajanga B, Kalluvya S, et al. Hospitalization outcomes in people living with HIV on Dolutegravir-based regimen in Mwanza, Tanzania: a comparative cohort. AIDS Res Ther [Internet]. 2025;22(1):11. Available from: https://aidsrestherapy.biomedcentral.com/articles/10.1186/s12981-025-00706-y

23. Reis KG, Desderius B, Kingery J, Kirabo A, Makubi A, Myalla C, et al. Blood pressure, T cells, and mortality in people with HIV in Tanzania during the first 2 years of antiretroviral therapy. J Clin Hypertens. 2020;22(9):1554–62.

24. Bantie B, Gebeyehu NA, Adella GA, Kassie GA, Mengstie MA, Abebe EC, et al. Trends of Body Mass Index changes among adults on antiretroviral therapy in Northwest Ethiopia: a longitudinal data analysis. Sci Rep [Internet]. 2024;14(1):1–11. Available from: 10.1038/s41598-024-53701-0

25. Sax PE, Erlandson KM, Lake JE, Mccomsey GA, Orkin C, Esser S, et al. Weight Gain Following Initiation of Antiretroviral Therapy: Risk Factors in Randomized Comparative Clinical Trials. 2020;71:1379–89.

26. Nyein PP, Petoumenos K, Borok M, Eriobu N, Kumarasamy N, Avihingsanon A, et al. Associations Between Antiretroviral Regimen and Changes in Blood Pressure: Results From the D2EFT Study. Clin Infect Dis [Internet]. 2024;80(1):160–3. Available from: 10.1093/cid/ciae256

27. Sempere A, Assoumou L, González-Cordón A, Waters L, Rusconi S, Domingo P, et al. Incidence of Hypertension and Blood Pressure Changes in Persons With Human Immunodeficiency Virus at High Risk for Cardiovascular Disease Switching From Boosted Protease Inhibitors to Dolutegravir: A Post-hoc Analysis of the 96-week Randomised NEAT-022 Tr. Clin Infect Dis [Internet]. 2023;77(7):991–1009. Available from: 10.1093/cid/ciad297

